# High-resolution single nucleotide polymorphisms detect chronological recombination events during Mpox Ib outbreak

**DOI:** 10.1101/2025.08.11.25333454

**Authors:** Michael C. Feehley, Patrick J. Feehley, Alex T. Poyer, Zih-Yu Hsieh, Gregory P. Contreras, Ting-Yu Yeh

**Author notes:** Corresponding author Ting-Yu Yeh, M.D., **Email:**. These authors contributed equally (Michael C. Feehley, Patrick J. Feehley). **Author Contributions:** MC Feehley, PJ Feehley, Contreras, and Yeh had full access to all the data in the study and take responsibility for the integrity of the data and the accuracy of the data analysis. Concept and design: Yeh, Michael and Patrick Feehley Acquisition, analysis, and interpretation of data: MC Feehley, PJ Feehley, and Yeh Statistical analysis and software: Poyer and Hsieh Edit: Yeh and Contreras. **Competing Interest Statement:** TYY is the CSO of Auxergen, Inc. and Auxergen s.r.l. GPC is the CEO of Auxergen, Inc. and Auxergen s.r.l. All authors declare no competing interests.

## Abstract

Viral recombination can rapidly generate new variants and influence outbreak severity. Currently, a method to track viral recombination events at high genomic resolution during natural transmission remains unavailable. Here we describe a novel linkage disequilibrium method and the first report of viral recombination events over time by using single nucleotide polymorphisms (SNPs) as natural genetic markers. Combined with phylogenetic data, we analyzed mpox virus (MPXV) Ib genome during 2023-2025 outbreak and found that 71 recombination events among Ib genomes, yielding a recombination frequency (Rf) and rate of 0.16%/kbp and 0.11%/kbp/year, respectively. Phylogenetic time analysis also shows that recombination began as early as December 23, 2023. Rf of MPXV Ib is 8.8-fold lower compared to vaccinia virus (1.5%/kbp) *in vitro*. Recombination and linkage hotspots are enriched at both inverted terminal repeat and variable regions of MPXV Ib genome. Our data not only reports the first Rf and rate of natural human transmission in the poxvirus family but also provides a powerful genomic surveillance tool for better understanding transmission and evolution of any viral outbreak globally.

## Introduction

Recombination is a significant driver of viral evolution and “;large-scale”; genetic changes that alter how viruses interact with their hosts (1, 2). When sequences contain high homology, homologous recombination is most common when replication machinery switches viral strain templates to create a new hybrid genome. While many focus on mutations as the greatest source of genetic variation in viruses, analyzing recombination help identify more long-term trends in viral adaptation and evolution over time, with the rise of new strains and resistance due to “;genetic conversion.”; Recombination increases favorable genetic diversity in viral populations as the quickest way to create new combinations of existing mutations.

The poxvirus family, particularly vaccinia virus, has long been one dsDNA virus of focus in recombination *in vitro* (3). However, the low recombination rates, a relatively large genome, and complex and repetitive regions in the poxvirus genome make ascertaining true crossovers at single nucleotide resolution a real challenge. To date, the recombination frequency (Rf) and rate during natural transmission of the poxvirus remains unknown. Due to the first-time widespread sequencing data available for mpox virus (MPXV) outbreak (4, 5), we develop a novel method to monitor MPXV Ib recombination events over time by using single nucleotide polymorphisms (SNPs) as natural genetic markers. This is the first report of natural Rf and recombination rate for the poxvirus family. Our results also reenforce that genomic surveillance is critical for understanding transmission and evolution of various viral outbreaks.

## Results and Discussion

While we and others reported recombination in the MPXV Ib and IIb outbreaks, the number of unique recombination events remained difficult to identify (4, 5, 7) as a disproportionate number of sequences were progeny of recombinants. One major obstacle to current SNP-based methods is difficulty in detecting low frequency SNPs, lack of genomics data, as well as complex patterns of coinfection and recombination (8). We therefore examined 213 MPXV sequences (October 13, 2023 to May 3, 2025) from Global Initiative on Sharing Influenza Data (GISAID) (Supplementary Material, Fig. S1). Using linkage disequilibrium (LD) analysis, we identified 450,545 total SNP pairs, with 1,602 pairs in strong linkage and 8,587 pairs as recombinants (Fig. 1A, Table S1) through Haploview (9), a significant frequency of detectable SNPs. Our method of SNP detection relies on changes in the consensus sequence of our Multiple Sequence Alignment (MSA) without an outgroup sequence, which allows accounting for low frequency differences along the raw alignment itself. While prior studies utilize individual SNPs per genome (10), we determined that analyzing SNP pairings under LD to be more resilient and more specific to track SNP phylogeny over complex evolution; through this, we identified 11 groups of shared recombinant SNP pairings across 213 sequences (Figue S2 and S3, Table S3). The quality of SNP pairing is very high, as evidenced by the fact that strong linkage SNP pair numbers are nearly half of the numbers of recombinant SNP pairs of recombinant MPXV sequences (R^2^ = 0.97, Figure S4). After associating phylogenetic time (branch length, IQ-TREE) with the number of SNP pairs (Fig. 1B and 1C, red), it suggested that the first recombination event occurred as early as December 23, 2023.

**Figure 1.**
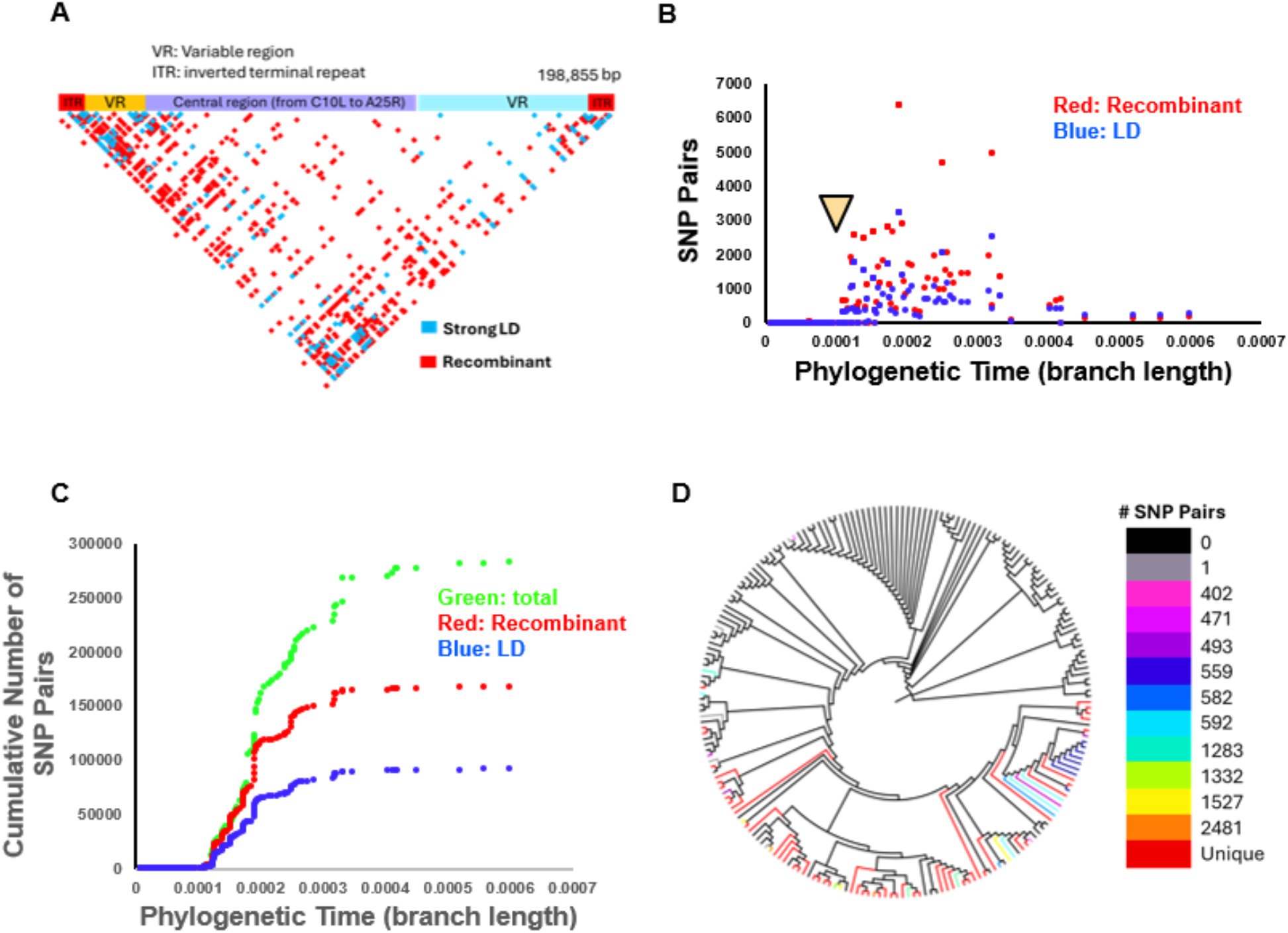
(A) Plot of recombinant and strong LD SNP pairs of 213 MPXV Ib sequences collected from October 13, 2023 to May 3, 2025. Confidence intervals are used to define strong linkage (CIHigh>0.98, CIlow>0.7) and recombinant (CIHigh<0.9). SNP pairs were then plotted with respect to their nucleotide positions along the MPXV Ib genome with SNP information provided by Haploview. (B) Plot of divergence and recombinant (red) and strong linkage (blue) SNP pairs. Python scripting associated phylogenetic time to each sequence’s SNP genotypes. Arrowhead labels the initiation date of recombination (December 23, 2023). (Table S1) (C) SNP data was supplemented with phylogenetic-based time; sequence ID and phylogenetic divergence (branch length, IQ-Tree) were assigned through custom Python scripting. Cumulative growth rate of total (green), recombinant (red) and strong linkage (blue) SNP pairs over phylogenetic time (Table S6). (D) Phylogenetic tree is graphed using FigTree and labeled by groupings of common recombinant SNP pair counts (by color, Table S3).

In our previous reports (4, 5), the recombination frequency of IIb or Ib remained unknown as accumulation of recombinant SNP pairs in lineages reduced the ability to identify unique recombination events. In fact, poxvirus recombination in natural transmission has never been reported to date. Using time analysis of SNP pairs, we discovered 89 sequences (41.7%) containing recombinant SNPs (Table S3). Unique recombination events can be distinguished from their progenies based on whether there is a change in recombination SNP pairs in the phylogenetic tree (Figure 1D). We identified 71 recombination events and their 18 progenies, yielding a recombination frequency (Rf) and rate of 0.17%/kbp and 0.11%/kbp/year, respectively. This is the first report of natural recombination frequency and rate in the poxvirus family. Given that 71 viruses of 213 viruses in our data set (∼33%) are circulating as unique recombinants, we speculate that current sequencing data may likely underreport current transmission or that MPXV infection potentially presents with high viral load in infected individuals (11), which is the reason for such high levels of recombination seen.

In addition, recombinant and strong LD SNP pairs were then used to create a hotspot map (N= 326 hotspots, Fig. 2A; and N=151 hotspots, Fig. 2B), showing enriched recombination and linkage at the variable regions and inverted terminal repeats; 20 haploblock breakpoints at recombination hotspots are also observed (Table S1). We found that 64 MPXV genes located in recombination hotspots (Table S4) and 32 genes contain strong linkage hotspots (Table S5) (32% and 16% of ∼200 MPXV total genes, respectively), suggesting large trends in inheritance and recombination. Among 151 strong LD hotspots, OPG172, and OPG166 contain the most linked SNPs. OPG153, OPG143, and OPG15 contain the most recombinant SNPs out of 326 recombination hotspots. The relationships between LD or recombinant hotspots that may change clinical outcomes and transmission of mpox outbreak require further investigation.

**Figure 2.**
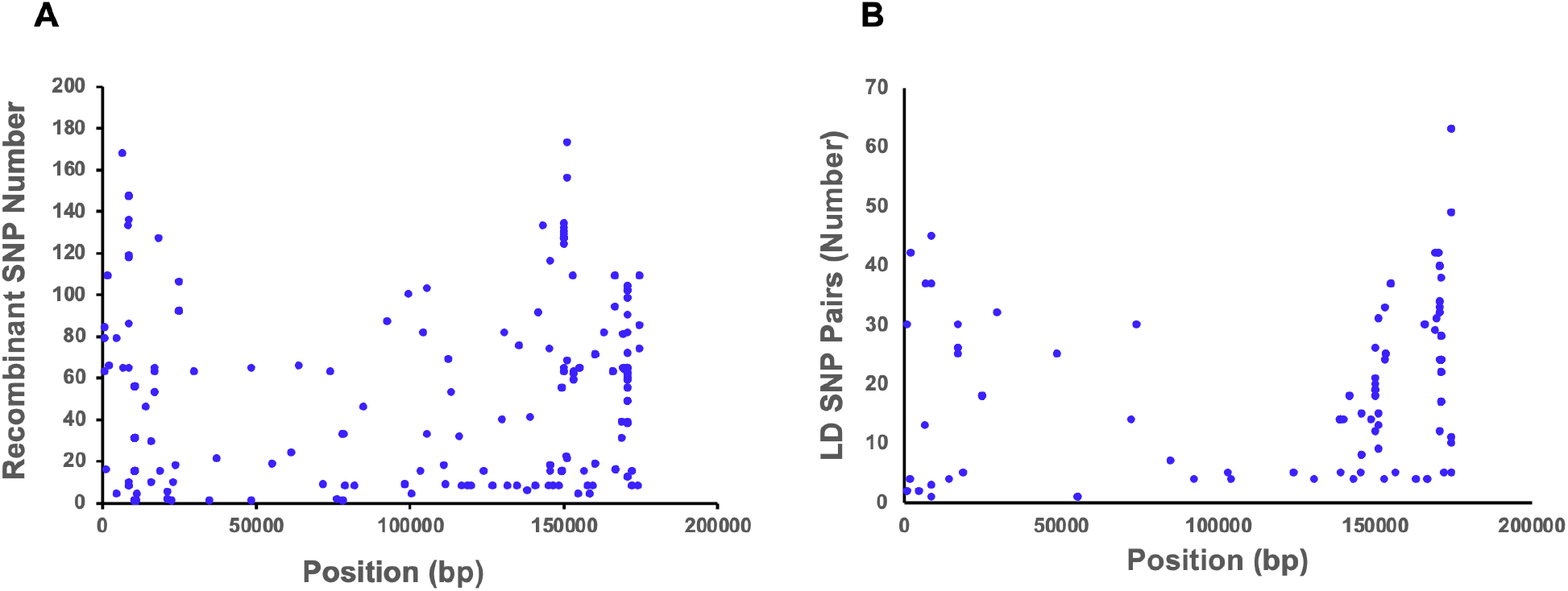
(A) Recombination hotspot map with recombinant SNP pairs per position are shown along the MPXV Ib genome of 213 sequences. Variables regions denoted under approximately 40 kbp endcaps with position information from 213 sequences. (B) Linkage hotspot map of strong linkage SNP pairs per position.

During our analysis, we discovered that Vero cell culture sequences (e.g. hMpxV/India/KL-ICMR-176_1073_P3/2024|EPI_ISL_19621542|2024-09-21) provide highly varied recombinant SNP profiles (6,659 more recombinant SNPs) compared to their clinical samples, suggesting that viral sequences cultured *in vitro* mutated differently than in natural transmission. Therefore, we did not include these cell culture sequences in this study. Natural MPXV Ib Rf is 8.8-fold lower than vaccinia virus (1.5%/kbp) *in vitro* (6). While a highly sensitive method especially for sequences with many recombinant SNP pairs, our method assumes a single recombination event each change in recombinant SNP pairs. Previous studies show that MPXV can swap DNA multiple times (6). Because we cannot ascertain whether a change in recombinant SNP pair number or type is always attributable to a single event, our method likely calculates a minimum number of recombination events, providing a reliable baseline estimate of Rf and recombination rate.

Many markers in viral genomes have been shown to be unreliable due to lack of resolution and availability, only recently improved with Covid-19 pandemic sequencing advances (12). Recombinant SNPs are intrinsically resilient to viral instability because multiple polymorphisms at the same site are critical to confirm recombination, increasing resolution at higher baseline mutation rates. Taken together, our SNP pair-wise analysis through LD provides a powerful genomic surveillance method for better understanding transmission and evolution of any viral outbreak globally.

## Data Availability

All data produced in the present study are available upon reasonable request to the authors

## Supplementary Information

### Materials and methods

303 MPXV Ib sequences (October 13, 2023 to May 3, 2025) were downloaded from the Global Initiative on Sharing Influenza Data (GISAID) website with the exclude low coverage (>25% ambiguous base count) filter. Python scripting further removed 90 sequences with >2,000 ambiguous bases as a second quality control step to prevent SNP contamination with ambiguous bases.

The phylogenetic tree was constructed by MAFFT7 and visualized using FigTree^1^. IQ-Tree with bootstrap 1000 assigned divergence time (branch length) to each sequence^2^ in the Multiple Sequence Alignment (MSA). Haploview^3^ was implemented to export SNP pairwise linkage disequilibrium (LD) data as well as confidence interval (CI) upper bound (CIhigh) and lower bound (CIlow) values. Strong LD (CIhigh>0.98 and CIlow>0.7) and recombinant (CIhigh<0.9) SNP pairs were extracted through Haploview’s plot of the 95% confidence bounds for *D*’, the normalized form of the degree of nonrandom association of two alleles, *D*, by *D*_max_. (Figure S1). A second MSA was done through MAFFT7 to align the reference sequence (ON563414.3 Monkeypox virus isolate MPXV_USA_2022_MA001, complete genome) with the 213 MSA. Python scripting rematched MSA SNP positions from Haploview back to the unaligned reference to determine the genes containing the SNP recombination and linkage hotspots.

Python scripting was done to match sequence SNP genotypes with Haploview’s calculation of pairwise SNP linkage. Genotypes with two of the calculated recombinant SNPs in each associated pair were marked as sequence positive for that specific SNP pair. IQ-Tree time-based phylogeny assignments were then made for each sequence, and therefore, SNP pair (Figure S2). Unique recombinant SNP pair counts were then sorted, while sequences with identical recombinant SNP pair counts were grouped together (Figure S3). Viral sequences with identical recombinant SNP pair genotypes were identified as progeny from a single recombination event. Sequences in the group, which showed a change in single or multiple recombinant SNP pair identities, were determined to be the result of an additional recombination event. Recombination frequency was derived by recombinant events, calculated as

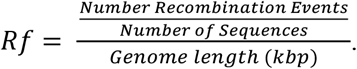

Resultant recombinant sequences (first recombinants) were identified by a change in color of the unrooted phylogenetic tree, which provides evolutionary directionality to the recombination events to determine when each event occurred with relation to each other. IQ-Tree phylogenetic time data was used to observe the distribution of SNP pairs over time to identify the first occurrence of a recombinant sequence by associating phylogenetic time to collection date.

## Supplementary Figures and Table Legends

**Figure S1.**
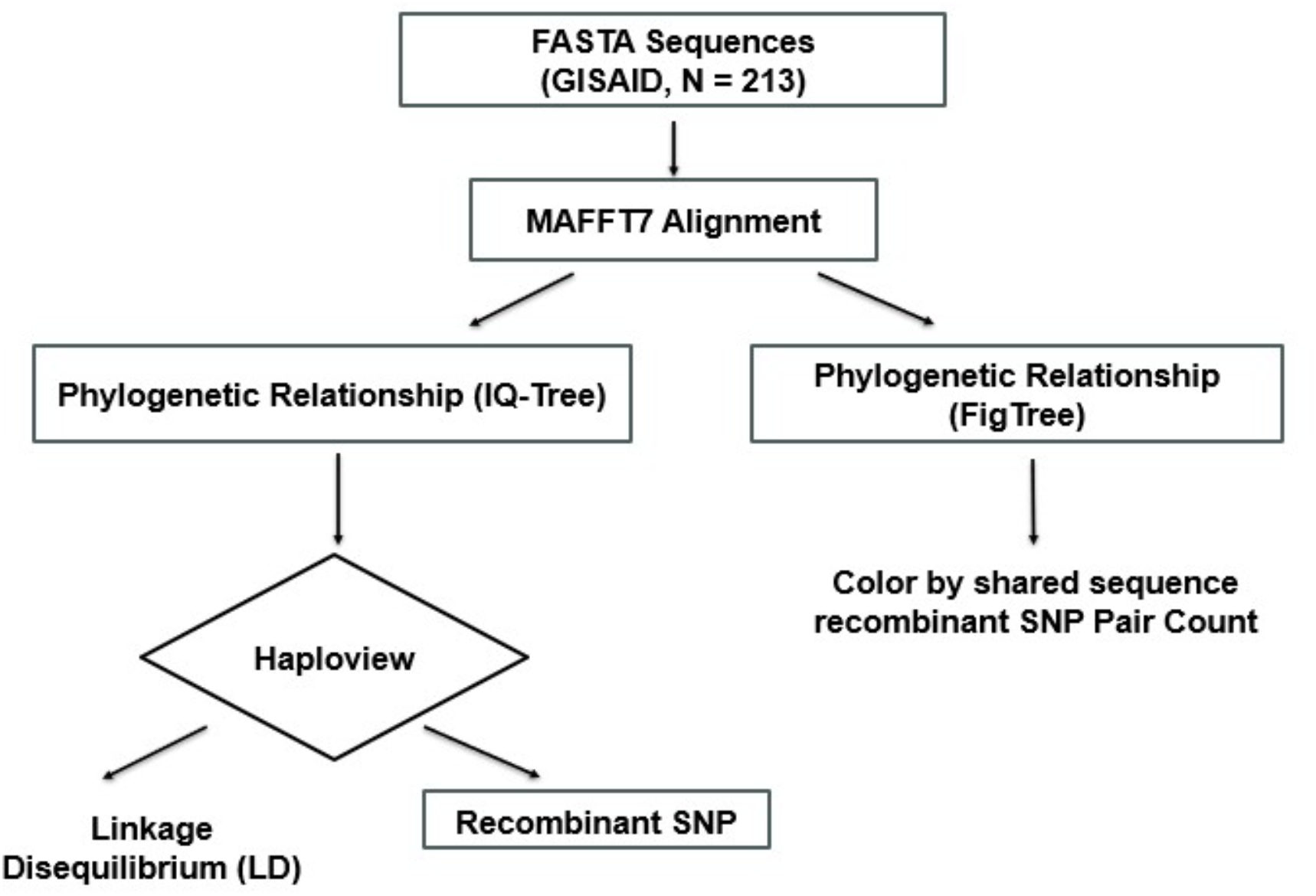
Flow chart of data processing in this study. Python scripting eliminated MPXV Ib sequences with > 2,000 base pairs of ambiguous bases; only human sample sequences were used (Oct 13, 2023 to May 3, 2025; N=213), while viral sequences from Vero cell cultures were excluded due to SNP outliers.

**Figure S2.**
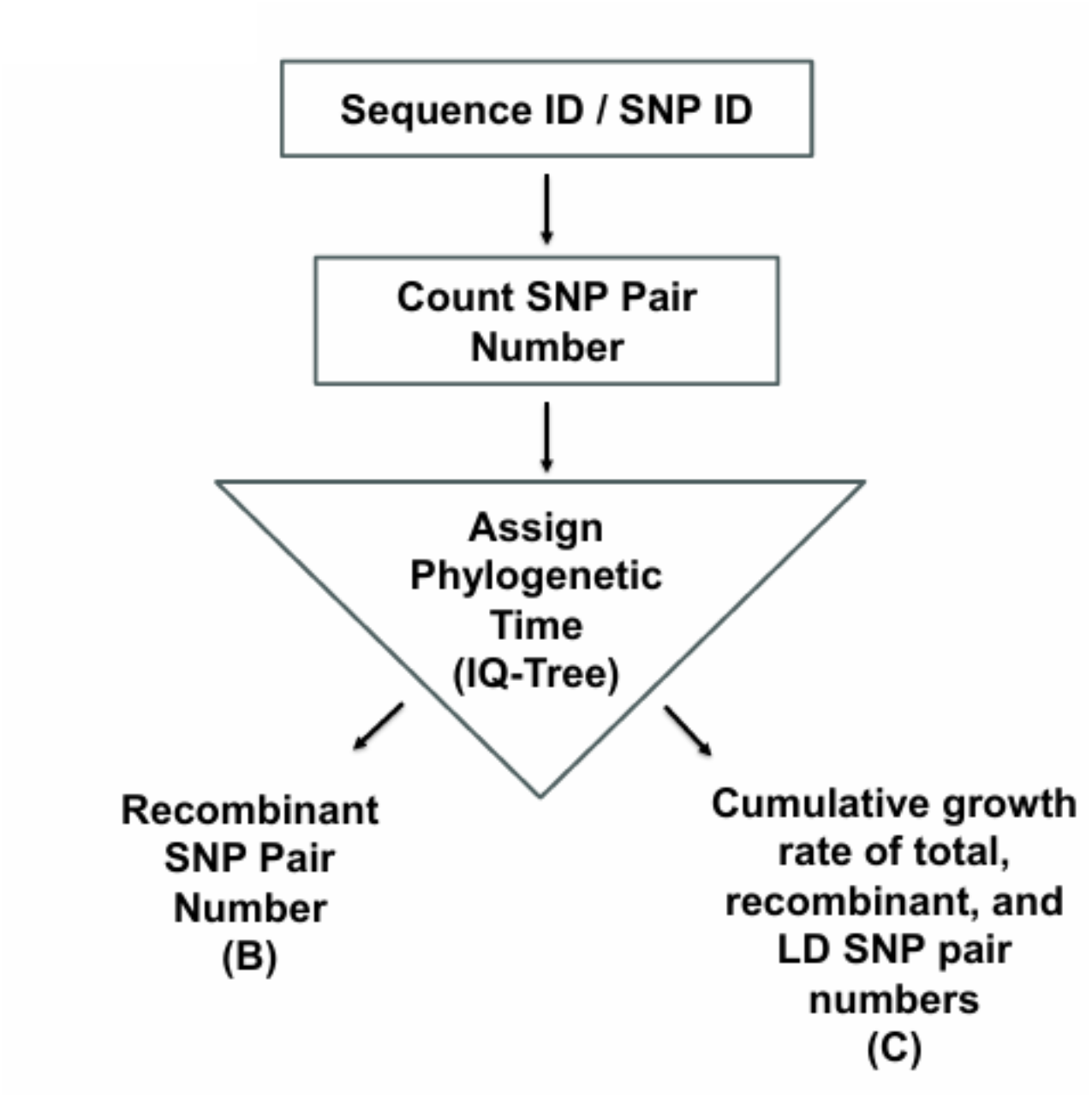
SNP data was supplemented with phylogenetic-based time; sequence ID and phylogenetic divergence (branch length, IQ-Tree) were assigned through Python scripting.

**Figure S3.**
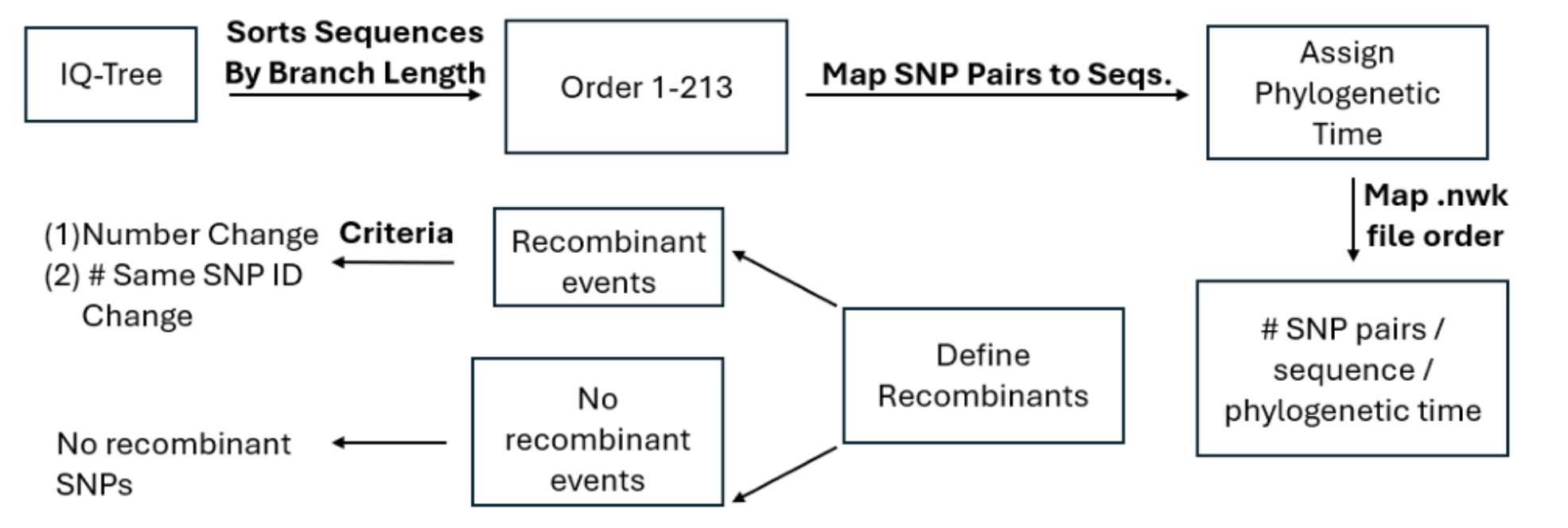
Phylogenetic data with SNP data was linked to sequence ID.

**Figure S4.**
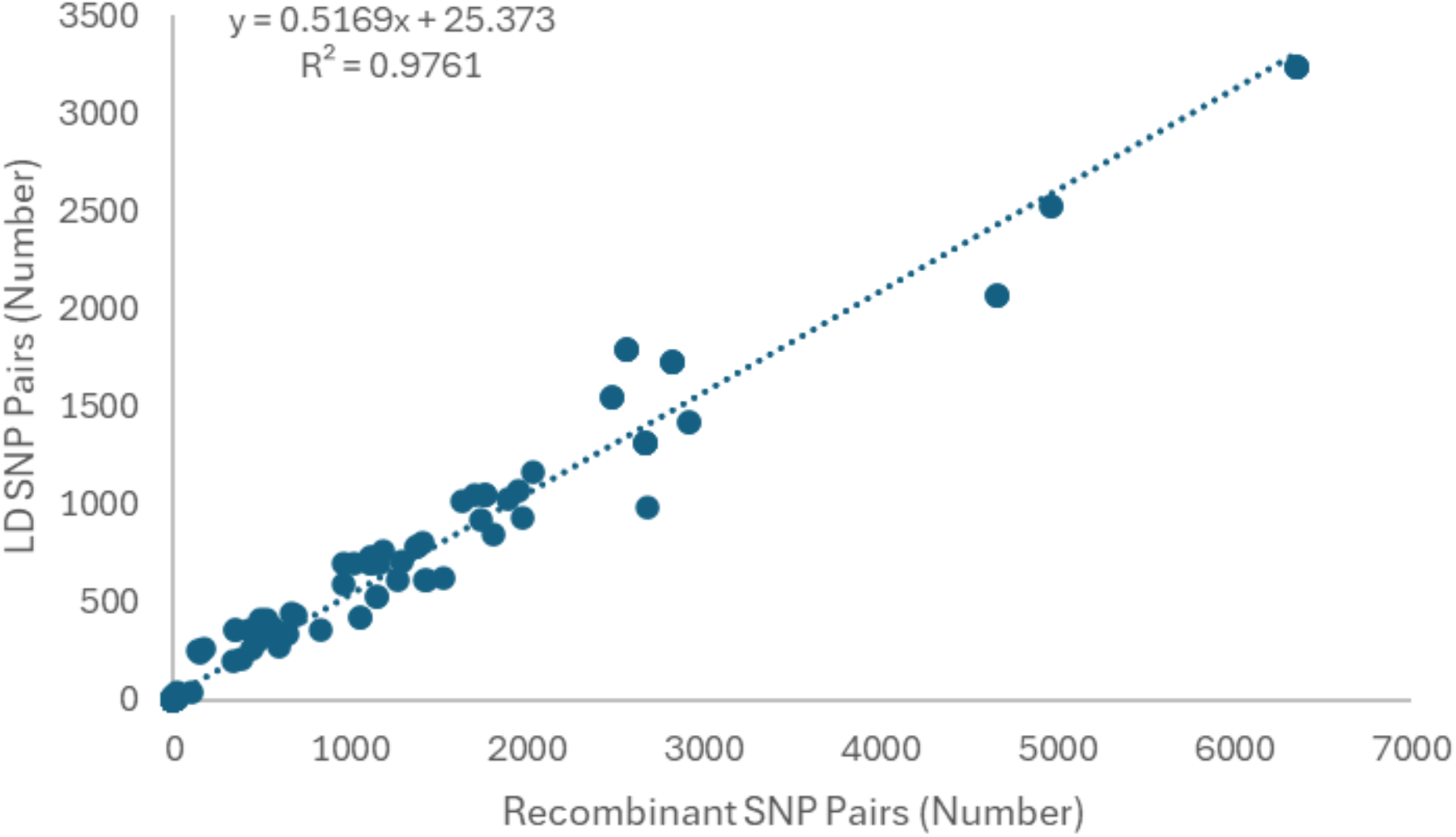
Plots of numbers of recombinant and strong linkage SNP pairs of 89 recombinant MPXV Ib sequences.

Table S1. Characterization of SNPs (per bp) that are recombination hotspots (326 hotspots total). Four gamete rule, solid spine, and Gabriel et al. methods defined haploblocks which were then processed to extract breakpoints. Table shows where breakpoints are also recombination hotspots (N = 20 for four gamete rule, solid spine, and the Gabriel et al. methods). LD and recombinant SNP pairs in this study are also listed with confidence interval (CI) upper bound (CIhigh) and lower bound (CIlow) values.

Table S2. Summary information of genomic mutations for all 213 MPXV Ib sequences in this study from GISAID.

Table S3. Identification of recombination event occurrences, their associated sequences, and characterization of progeny viruses. Color scheme of each sequence is shown in Figure 1D.

Table S4 and S5. Map of SNP positions from aligned and unaligned reference sequence (ON563414.3 Monkeypox virus isolate MPXV_USA_2022_MA001, complete genome) for recombinant SNPs and strong LD SNPs.

Table S6. Cumulative growth rate of total, recombinant and strong linkage SNP pairs over phylogenetic time (Figure 1C).

